## Supplementary information for "Species-level verification of *Phascolarctobacterium* association to colorectal cancer"

### Supplementary figures

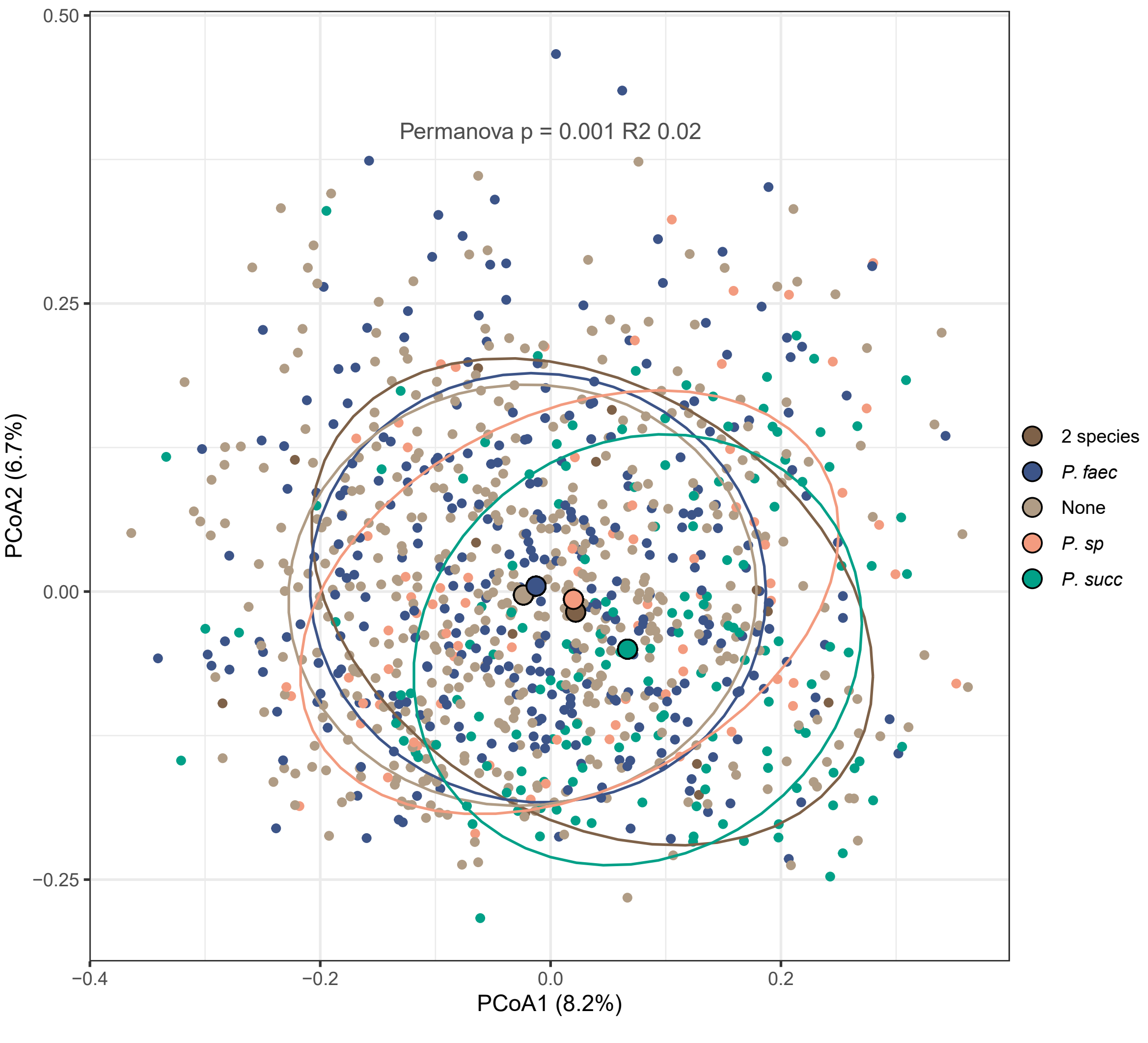


Figure S1: PCoA plot of microbiome composition in CRCbiome samples. Groups are defined as those samples that have reads from only *P. succinatutens*, *P.* sp 377, *P. faecium*, two *Phascolarctobacterium* or no *Phascolarctobacterium*. PERMANOVA results showed a significant difference between groups ($p<0.001$) with an $R^2$ of 0.02.

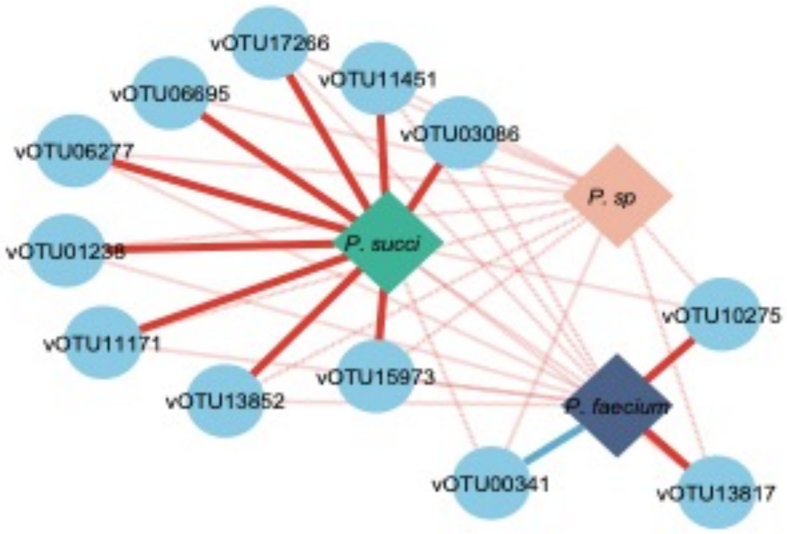


Figure S2: vOTU correlation plot. Presence/absence-based co-occurrence network between *Phascolarctobacterium* species and viruses in the CRCbiome dataset. Solid lines indicate significant correlations (FDR-corrected $p < 0.05$), whereas shaded lines indicate non-significant correlations. Only vOTUs with a significant correlation to one of the *Phascolarctobacterium* MAGs are depicted. Red: tendency to co-exclusion; Blue: tendency to co-existence. The direction of the correlation was defined based on the difference between the observed “yes/yes" and the expected “yes/yes”.
